# Access to COVID-19 Vaccines in High-, Middle-, and Low-Income Countries Hosting Clinical Trials

**DOI:** 10.1101/2021.07.16.21260509

**Authors:** Reshma Ramachandran, Joseph S. Ross, Jennifer E. Miller

## Abstract

The COVID-19 pandemic has led to the rapid development of multiple vaccines, vaccines that were tested in clinical trials located in several countries around the world. Because prior research has shown that pharmaceuticals do not receive consistent and timely authorization for use in lower-income countries where they are tested, we conducted a cross-sectional study examining the authorization or approval and delivery for COVID-19 vaccines recommended by the World Health Organization (WHO) in the countries where they were tested. While countries of varying incomes have largely authorized the COVID-19 vaccines tested within their populations for use, high-income countries have received proportionately more doses, enabling them to more fully vaccinate their populations. As many lower-income countries continue to experience inequitable shortfalls in COVID-19 vaccine supply amid the ongoing pandemic, efforts must be undertaken to ensure timely access in countries across all income groups, including those hosting clinical trials.

## Introduction

The COVID-19 pandemic has led to the rapid development of multiple vaccines, vaccines that were tested in clinical trials located in several countries around the world.^1^ However, low- and middle-income countries have experienced significant delays in vaccine access despite mechanisms meant to ensure fair distribution, such as COVAX.^2^ Because prior research has shown that pharmaceuticals do not receive consistent and timely authorization for use in lower income countries where they are tested,^3^ we examined the authorization or approval and delivery for COVID-19 vaccines recommended by the World Health Organization (WHO) in the countries where they were tested.

## Methods

For this cross-sectional study, we identified COVID-19 vaccines listed by the WHO for emergency use as of June 8, 2021. We then identified all completed clinical trials for these vaccines using the WHO COVID-19 Vaccine Tracker and Landscape and the McGill University COVID-19 Vaccine Tracker. We then extracted trial primary completion dates, phase, and country locations from ClinicalTrials.gov. Each country was classified by national income group using the World Bank historical classifications for this year (2021).^4^

Through regional and national regulatory agency websites, we determined whether these countries hosting vaccine clinical trials also authorized or approved its use (hereafter, authorized). Additionally, for each country, we extracted data on doses procured and delivered from the UNICEF COVID-19 Vaccine Market Dashboard and the Airfinity COVID-19 platform. We calculated the proportion of countries hosting clinical trials authorizing any vaccine, and the vaccine tested in their population, and their delivery, overall and for individual manufacturers. Lastly, we determined the median proportion of people aged 15 years and older in each country able to receive a full vaccination series using population data from the United Nation’s World Population Dashboard. We conducted descriptive statistical analyses, estimating medians and proportions, using Excel spreadsheet software version 16.0 (Microsoft).

## Results

The six unique COVID-19 vaccines, listed for emergency use by the WHO as of June 8, 2021, were tested in 21 countries (Table 1). Among 10 high-income countries hosting completed clinical trials, 9 (90.0%) authorized the tested vaccine and received enough doses to vaccinate a median 12.4% (IQR,1.6-24.4) of these countries’ populations aged 15 years and older. Among the 10 upper-middle income countries, rates were 90.0% and 3.4% (IQR,0.8-19.6), respectively. Examining ongoing and completed trials, corresponding rates were higher for high-income countries, but lower for upper middle-income countries (Table 1).

**Table 1.** Access to any COVID-19 vaccine in countries with completed clinical trials categorized by national income group (as of June 8, 2021)

| Countries Tested<br>(World Bank<br>Classification) | Countries with<br>Clinical Trials,<br>No. (%) | Countries with Clinical<br>Trials with<br>Authorization/Approval,<br>No. (%) | Countries with<br>Clinical Trials with<br>Delivered Doses,<br>No. (%) | Vaccination Series<br>Delivered Per Capita<sup>a</sup> in<br>Countries with Clinical<br>Trials, Median % (IQR) |
| --- | --- | --- | --- | --- |
| **Completed Clinical**<br>**Trials** | 21 | 19 (90.5) | 18 (85.7) | 7.1 (1.4-23.7) |
| Low-Income | 0 (0.0) | N/A | N/A | N/A |
| Lower Middle-Income | 1 (4.8) | 1 (100.0) | 1 (100.0) | 11.2 |
| Upper Middle-Income | 10 (47.6) | 9 (90.0) | 8 (80.0) | 3.4 (0.8-19.6) |
| High-Income | 10 (47.6) | 9 (90.0) | 9 (90.0) | 12.4 (1.6-24.4) |
| **Ongoing and**<br>**Completed Clinical**<br>**Trials** | 32 | 29 (90.6) | 29 (90.6) | 15.0 (2.1-26.5) |
| Low-Income | 0 (0.0) | N/A | N/A | N/A |
| Lower Middle-Income | 4 (12.5) | 4 (100.0) | 4 (100.0) | 6.4 (3.5-10.3) |
| Upper Middle-Income | 13 (40.6) | 10 (76.9) | 10 (76.9) | 2.5 (1.1-16.9) |
| High-Income | 15 (46.9) | 15 (100.0) | 15 (100.0) | 29.8 (19.6-38.8) |

<sup>a</sup> Data at the country level for populations ages 15-64 as well as 65 and above was extracted from the World Population Dashboard hosted by the United Nations Population Fund. Data for populations ages 18 and above as well as other specific age groups was unable to be extracted.

While Moderna completed all clinical trials in one country, AstraZeneca and Janssen completed trials in 10 and 12 countries, respectively (Table 2), receiving authorization for their vaccines in 60.0% and 75.0% of these countries, respectively. Across manufacturers, high-income countries received more doses to vaccinate larger median proportions of countries’ populations aged 15 years and older. Delivery of doses procured by COVAX ranged between 0% and 9.8% with no doses delivered of Moderna or Janssen vaccines as of June 8, 2021.

**Table 2.** Access to COVID-19 vaccines in countries hosting completed clinical trials categorized by manufacturer (as of June 8, 2021)

| Manufacturer (Vaccine) | Number of Countries with Completed Clinical Trials | Countries with Completed Clinical Trials with Authorization/Approval, No. (%) | Countries with Completed Clinical Trials with Doses Delivered, No. (%) | Completed Trials in LMICs (Median % Delivered Per Capita<sup>b</sup>, range) | Completed Trials in UMICs (Median % Delivered Per Capita<sup>b</sup>, range) | Completed Trials in HICs (Median % Delivered Per Capita<sup>b</sup>, range) | COVAX Doses Procured (% Delivered) |
| --- | --- | --- | --- | --- | --- | --- | --- |
| Pfizer-BioNTech (BNT162b2) | 6 | 6 (100.0) | 5 (83.3) | 0 (N/A) | 4 (0.7, 0.0-2.2) | 2 (29.8, 24.7-34.9) | 40,000,000 (6.8) |
| Moderna (mRNA-1273) | 1 | 1 (100.0) | 1 (100.0) | 0 (N/A) | 0 (N/A) | 1 (27.3) | 500,000,000 (0.0) |
| Janssen (Ad26.COV2.S) | 12 | 10 (83.0) | 5 (41.7) | 0 (N/A) | 6 (0.0, 0.0-0.4) | 6 (1.0, 0.0-7.6) | 500,000,000 (0.0) |
| AstraZeneca/Serum Institute of India (AZD1222/Vaxzevria/Covishield)<sup>a</sup> | 10 | 7 (70.0) | 8 (80.0) | 0 (N/A) | 6 (1.3, 0.0-10.4) | 4 (3.8, 0.0-36.5) | 720,000,000 (9.8) |
| Sinopharm (BBIBP-CorV) | 5 | 5 (100.0) | 5 (100.0) | 1 (7.4) | 2 (9.8, 7.1-12.4) | 2 (19.6, 17.6-21.5) | 0 |
| Sinovac (CoronaVac) | 3 | 3 (100.0) | 3 (100.0) | 0 (N/A) | 3 (13.5, 12.2-21.6) | 0 (N/A) | 0 |

<sup>a</sup>As the same efficacy data was submitted for the vaccine Covishield, manufactured by the Serum Institute of India as the vaccine AZD1222 (Vaxzevria) by their common developers, Oxford University and AstraZeneca, this was considered to be a single vaccine for the analyses. <sup>b</sup>This refers to the population within tested countries that is 15 years and older.

## Discussion

While countries of varying incomes have largely authorized the COVID-19 vaccines tested within their populations for use, high-income countries have received proportionately more doses, enabling them to more fully vaccinate their populations. These findings parallel ongoing access disparities, as high-income countries have successfully procured^5^ and administered^6^ doses ahead of low- and middle-income countries.

Study limitations include inability to account for the number of participants enrolled in vaccine trials per country, which was not systematically reported for all trials, and inability to account for the impact of manufacturing errors and safety concerns on dosage delivery, which particularly affected AstraZeneca and Janssen.

As many lower-income countries continue to experience inequitable shortfalls in COVID-19 vaccine supply amid the ongoing pandemic, efforts must be undertaken to ensure timely access in countries across all income groups, including those hosting clinical trials.

## Data Availability

Dr. Ramachandran had full access to all the data in the study and take responsibility for the integrity of the data and the accuracy of the data analysis. Requests for the dataset can be made to the corresponding author at.

## Data access and responsibility

Dr. Ramachandran had full access to all the data in the study and take responsibility for the integrity of the data and the accuracy of the data analysis.

## Author contributions

RR, JSR, and JEM contributed to study concept and design; RR abstracted the data; all authors contributed to the analysis and interpretation of the data; RR drafted the manuscript; all authors contributed to the critical revision of the manuscript; and JEM provided study supervision.

## References

1. World Health Organization. COVID-19 vaccine tracker and landscape. Accessed June 25, 2021. https://www.who.int/publications/m/item/draft-landscape-of-covid-19-candidate-vaccines

2. Usher AD. A beautiful idea: how COVAX has fallen short. The Lancet. 2021;397(10292):2322–2325. doi:10.1016/S0140-6736(21)01367-2

3. Miller JE, Mello MM, Wallach JD, et al. Evaluation of Drug Trials in High-, Middle-, and Low-Income Countries and Local Commercial Availability of Newly Approved Drugs. JAMA Netw Open. 2021;4(5):e217075. doi:10.1001/jamanetworkopen.2021.7075

4. World Bank Data Help Desk. World Bank Country and Lending Groups. Accessed June 25, 2021. https://datahelpdesk.worldbank.org/knowledgebase/articles/906519-world-bank-country-and-lending-groups

5. So AD, Woo J. Reserving coronavirus disease 2019 vaccines for global access: cross sectional analysis. BMJ. 2020;371:m4750. doi:10.1136/bmj.m4750

6. Cohen J, 2020, Pm 7:30. Early approval of a COVID-19 vaccine could stymie the hunt for better ones. Science | AAAS. Published online October 14, 2020. https://www.sciencemag.org/news/2020/10/early-approval-covid-19-vaccine-could-stymie-hunt-better-ones

